# Cardiac and Perinatal Outcomes Among Pregnancies with Maternal Cardiac Arrhythmias

**DOI:** 10.1101/2025.10.09.25337697

**Authors:** Gianna Wilkie, Sruthi Takillapati, Majd Al Deen Alhuarrat, Anna Whelan, Lawrence Rosenthal, Lara C. Kovell

## Abstract

**Background:** Atrial fibrillation/flutter (AF/AFL) and supraventricular tachycardia (SVT) are common arrhythmias in pregnancy. While known to contribute to adverse outcomes, large studies have not evaluated cardiac and perinatal outcomes in individuals with these arrhythmias.

**Objective:** To characterize the cardiac/perinatal outcomes of patients with AF/AFL and SVT in pregnancy.

**Methods:** We performed a retrospective cohort study utilizing Epic Cosmos, a dataset from over 1,500 US hospitals. We identified pregnancies between October 25, 2021 and October 24, 2024 of patients with a diagnosis of AF/AFL and SVT using simultaneous diagnoses codes for each arrhythmia and pregnancy. A control group of pregnancies without arrhythmias was also identified. Descriptive statistics were calculated, and comparisons were made with ANOVA or Chi-square tests, as appropriate.

**Results:** During the study period, there were 8,642 pregnant patients with AF/AFL, 29,638 with SVT, and 5,372,171 healthy pregnant controls. Patients with these arrhythmias were older (AF/AFL: 33±6, SVT: 32±6, control: 31±6, *p*<0.0001) with a higher BMI than the control group. Those with arrhythmias experienced higher rates of heart failure (AF/AFL: 10.8%, SVT: 3.8%, control: 0.3%), thromboembolism, stroke, preterm birth, and cesarean delivery (all *p*<0.0001). Patients with AF/AFL had 23.9 times higher odds of stroke compared to the control group (95% CI 21.3-27.0).

**Conclusions:** While both AF/AFL and SVT are associated with adverse cardiac and perinatal outcomes, AF/AFL was associated with the highest risk of heart failure and stroke. Further research is needed to understand optimal perinatal AF/AFL management, especially anticoagulation therapy given the high rate of maternal stroke.

## Introduction

Cardiovascular disease is the leading cause of maternal mortality in pregnancy and impacts around 1 to 4% of pregnancies in the United States.^1,2^ Arrhythmia is a growing complication seen among pregnant patients that can predispose the maternal-child dyad to severe adverse outcomes.^3^ Current characteristics of the maternal population contributing to an increased risk of arrhythmias in pregnancy include a rising average maternal age, widespread and increasing diabetes mellitus and obesity, and a growing number of individuals with congenital heart disease (CHD) surviving to their reproductive years.^2^

Arrhythmias that arise in pregnancy are mostly self-limiting and resolve postpartum, but not all are benign.^4^ Commonly identified arrhythmias in pregnancy are atrial fibrillation (AF)/atrial flutter (AFL) and supraventricular tachycardia (SVT), which can occur in pregnancy for the first time or worsen previously well controlled arrhythmias.^5^ Review articles suggest that atrial fibrillation is associated with an increase in maternal mortality, but also note limited information on the risk of stroke in pregnancy in this high-risk population.^6,7^ Expert consensus suggests the decision to provide treatment depends on the severity of the arrhythmia and the risks and benefits to the mother and fetus.^4^ However, outside of the CHA2DS2-VASc score, there is minimal current data and pregnancy-specific clinical guidelines available to support the increasing need of clinical decision making in pregnancies complicated by arrhythmias like AF/AFL and SVT.^4^

The objective of this study is to characterize the frequency of cardiac arrhythmias in pregnancy in a large, modern cohort, and analyze the effects of AF/AFL and SVT during pregnancy on perinatal and cardiac outcomes.

## Methods

We conducted a retrospective cohort study of pregnancies and patients from the Epic Cosmos database, a comprehensive electronic health care record (EHR) database encompassing data from over 1,704 hospitals across the United States, D.C., Lebanon, and Saudi Arabia. This database represents more than 296 million patients with participating healthcare systems voluntarily contributing a limited dataset, which is centrally anonymized by Epic. Due to the publicly available/de-identified nature of this data, this study was deemed to be exempt from the institutional review board at the UMass Chan School of Medicine. This study adheres to Strengthening the Reporting of Observational Studies in Epidemiology (STROBE) guidelines.^8^

We queried the most recently available data at the time of our study from October 25, 2021, through October 24, 2024 identifying our base population of pregnancies. All pregnancies were then linked to patients with overlapping ICD-10-CM codes of AF/AFL (I48.*) and SVT (I47.1*). A separate cohort without a history of arrhythmia utilizing the Epic Cosmos ACC grouper for arrhythmia was built by excluding this grouper.

Aggregate measures of demographics and body measures collected include age, race/ethnicity (self-reported in Epic), and body mass index (BMI). Patients may report multiple races in Epic. CHA_2_DS_2_-VASc scores were also abstracted from Epic Cosmos, and patients may have had multiple documented scores over the included time period due to changing risk factors. Cardiac medications including beta-blockers, calcium channel blockers, anti-arrhythmic medications (including sodium and potassium channel blockers), and anti-coagulant medications were reviewed. Cardiac outcomes included stroke, thromboembolism, heart failure, and endocarditis. Patients were then linked to pregnancies to assess all perinatal outcomes, and patients may have had multiple pregnancies during the time frame. Perinatal outcomes included mean gestational age at delivery, development of gestational diabetes mellitus or pre-eclampsia, term delivery (≥37 weeks of gestational age), preterm birth (less than 37 weeks), pregnancy loss or termination at less than 22 weeks of gestation, and cesarean delivery.

Descriptive statistics with raw numbers and percentages or mean and standard deviation were reported for all aggregate demographics and outcomes. Comparisons between those with AF/AFL, SVT, and a control group of pregnant patients without an arrhythmia diagnosis were made with ANOVA for continuous variables or Chi-square test for categorical variables where appropriate. A total of 16 Chi-square or ANOVA tests were performed to assess group differences. To control for type I error due to multiple comparisons, a Bonferroni correction was applied. The original alpha level of 0.05 was divided by the number of tests, resulting in an adjusted significant p value of <0.003. Pairwise comparisons were then made between those with either AF/AFL or SVT and the control group with no arrhythmia using *t*-test or Chi-square with the same Bonferroni correction (p<0.003 deemed significant). No imputation was performed for missing CHA_2_DS_2_-VASc scores, but those missing CHA_2_DS_2_-VASc scores were excluded from score-specific analyses. An unadjusted odds ratio was also calculated for stroke and heart failure by type of arrhythmia exposure. Due to the design of the dataset, if less than 10 individuals were identified to have an outcome, the exact number was not provided to maintain patient confidentiality.

## Results

During the study period, there were 8,642 pregnant patients with AF/AFL, 29,638 with SVT, and 5,372,171 pregnant controls with no history of arrhythmia. There were a total of 6,468,084 pregnancies included in the total cohort, of which 38,280 (0.6%) were affected by AF/AFL or SVT.

There were statistically significant differences in age, race, ethnicity, and BMI between pregnant patients with AF/AFL, SVT, and no arrhythmias. Pregnant patients with AF/AFL were a mean age of 33 (±6) years old, 66.4% White, and 84.1% non-Hispanic, while pregnant patients with SVT were a mean age of 32 (±6) years old, 80.0% White, and 86.6% non-Hispanic (Table 1). There were also statistically significant differences in medical history including pregestational diabetes mellitus, chronic hypertension, chronic kidney disease, cardiomyopathy, and congenital heart disease between these three groups. Nearly half of patients with AF/AFL had chronic hypertension (47.5%) and 9.6% had pre-gestational diabetes mellitus, whereas 36.7% of patients with SVT had chronic hypertension and 5.1% had pre-gestational diabetes mellitus. The CHA_2_DS_2_-VASc score of patients with AF had significant missing data (n=7,808). Of those with a score reported, 767 (91.9%) patients had a score of 1, 366 (43.9%) had a score of 2, 108 (12.9%) had a score of 3, 41 (4.9%) had a score of 4, and 16 (1.9%) had a score of 5 or more. Additionally, 464 (55.6%) patients with a documented score had multiple CHA_2_DS_2_-VASc scores available over the study time period.

**Table 1.** Demographics of Pregnant Patients with Cardiac Arrhythmias.

| Demographics | Pregnant Patients with Atrial Fibrillation/Flutter (n= 8,642) | Pregnant Patients with SVT (n= 29,638) | Pregnant Patients with no Arrhythmia History (n=5,372,171) | p-value |
| --- | --- | --- | --- | --- |
| Age, years (mean ± SD) | 33 (±6) | 32 (±6) | 31 (±6) | <0.0001 |
| Race<sup>a</sup> |  |  |  |  |
| American Indian/Alaskan Native | 166 (1.9%) | 404 (1.4%) | 85,432 (1.6%) | <0.00001 |
| Asian | 281(3.3%) | 980 (3.3%) | 325,563 (6.0%) |  |
| Black | 2370 (27.4%) | 4,222 (14.2%) | 1,052,615 (19.6%) |  |
| Native Hawaiian/Other Pacific Islander | 61 (0.7%) | 189 (0.6%) | 45,865 (0.9%) |  |
| White | 5,734 (66.4%) | 23,736 (80.0%) | 3,395,688 (63.2%) |  |
| Other | 1,333 (15.4%) | 3,898 (13.2%) | 971,627 (18.1%) |  |
| Unknown | 90 (1.0%) | 316 (1.1%) | 237,493 (4.4%) |  |
| Ethnicity |  |  |  |  |
| Hispanic | 921 (10.7%) | 2,611 (8.8%) | 1,181,772 (22.0%) | <0.0001 |
| Non-Hispanic | 7,271 (84.1%) | 25,669 (86.6%) | 3,829,229 (71.3%) |  |
| Unknown | 450 (5.2%) | 1,358 (4.6%) | 360,170 (6.7%) |  |
| Body Mass Index (kg/m<sup>2</sup>) | 33.1 (±11.3) | 31.1 (±7.0) | 31.7 (±10.1) | <0.0001 |
| History of Pre-gestational Diabetes | 831 (9.6%) | 1,523 (5.1%) | 137,410 (2.6%) | <0.0001 |
| History of Chronic Hypertension | 4,106 (47.5%) | 10,866 (36.7%) | 1,100,128 (20.5%) | <0.0001 |
| History of Chronic Kidney Disease | 289 (3.3%) | 385 (1.3%) | 17,859 (0.3%) | <0.0001 |
| Cardiomyopathy | 648 (7.5%) | 1,025 (3.5%) | 14,302 (0.3%) | <0.0001 |
| Congenital Heart Disease | 684 (7.9%) | 1,661 (5.6%) | 33,945 (0.6%) | <0.0001 |

All data are reported as n (%) or mean ( ± standard deviation). *P*-values are reported significant at $p<0.05$ for comparisons across all three groups.
<sup>a</sup>Subjects may identify with multiple races.

Patients with AF/AFL and SVT were significantly more likely to be prescribed cardiac medications during pregnancy compared to those without arrhythmias (Table 2). Beta blocker use was highest in the AF/AFL group (59.3%), followed by SVT (52.9%) and controls (12.6%, *p* < 0.0001), with overall lower use of calcium channel blockers. Use of sodium channel blockers and potassium channel blockers were also highest in the AF/AFL group (AF/AFL: 12.3% and 4.6% vs. SVT: 6.7% and 1.0%, respectively). Anticoagulant medications were used at any point in pregnancy by 58.7% of AF/AFL patients, 35.1% of SVT patients, and 16.3% of controls (*p* < 0.0001).

**Table 2.** Cardiac Outcomes of Pregnant Patients with Cardiac Arrhythmias.

| **Cardiac Outcomes** | **Pregnant Patients with Atrial Fibrillation/Flutter (n= 8,642)** | **Pregnant Patients with SVT (n= 29,638)** | **Pregnant Patients with no Arrhythmia History (n=5,372,171)** | ***p*-value** |
| --- | --- | --- | --- | --- |
| **Medications Used** |  |  |  |  |
| Beta Blockers | 5,121 (59.3%)‡ | 15,683 (52.9%)‡ | 679,139 (12.6%) | <0.0001 |
| Calcium Channel Blockers | 2,822 (32.7%)‡ | 6,394 (21.6%)‡ | 538,052 (10.0%) | <0.0001 |
| Sodium Channel Blocker/Antiarrhythmic | 1,061 (12.3%)‡ | 1,979 (6.7%)‡ | 77,940 (1.5%) | <0.0001 |
| Potassium Channel Blocker/Antiarrhythmic | 399 (4.6%)‡ | 306 (1.0%)‡ | 679 (0.01%) | <0.0001 |
| Anticoagulant Medications | 5,075 (58.7%)‡ | 10,401 (35.1%)‡ | 878,023 (16.3%) | <0.0001 |
| **Cardiac Outcomes** |  |  |  |  |
| **Heart Failure** | 930 (10.8%)‡ | 1,133 (3.8%)‡ | 15,325 (0.3%) | <0.0001 |
| **Thromboembolism** | 542 (6.3%)‡ | 909 (3.1%)‡ | 30,746 (0.6%) | <0.0001 |
| **Stroke** | 300 (3.5%)‡ | 306 (1.0%)‡ | 8,058 (0.1%) | <0.0001 |
| **Endocarditis** | 165 (1.9%)‡ | 211 (0.7%)‡ | 3,333 (0.06%) | <0.0001 |

All data are reported as n (%) or mean ( ± standard deviation). *P*-values are reported significant at $p<0.05$ in the table from ANOVA or Chi square comparison across all three groups.
‡ Denotes a significant difference in pairwise comparisons between atrial fibrillation/flutter and control group of no arrhythmia history or SVT and control group of no arrhythmia history (Bonferroni adjusted $p<0.003$ ).

Patients with these arrhythmias in pregnancy experienced more adverse cardiac outcomes compared to the healthy controls. Among pregnant patients with AF/AFL, 10.8% developed heart failure and 3.5% experienced a stroke, while in those with SVT 3.8% developed heart failure and 1.0% a stroke (Table 2). The unadjusted odds of heart failure among patients with AF/AFL when compared to those without arrhythmia was OR 42.2 (95% CI 39.3-45.2), while the odds of heart failure among patients with SVT as compared to those without arrhythmia was OR 13.9 (95% CI 13.1-14.8). The unadjusted odds of stroke among patients with AF/AFL when compared to those without arrhythmia was OR 23.9 (95% CI 21.3-27.0) while the odds of stroke among patients with SVT as compared to those without arrhythmia was OR 6.9 (95% CI 6.2-7.8) (Table 2).

In terms of perinatal outcomes, there were statistically significant differences in gestational age at delivery, development of pre-eclampsia and gestational diabetes mellitus, preterm birth, and mode of delivery between the three groups (Table 3). Approximately 16.2% of pregnant patients with AF/AFL delivered preterm as compared to 11.8% of pregnant patients with SVT and 8.1% of healthy controls (*p* < 0.0001). The rate of cesarean delivery was 33.7% among pregnancies with AF/AFL, 29.8% of pregnancies with SVT, and 23.8% with no arrhythmia history (p<0.0001). Pregnancies with AF/AFL were the most likely to develop pre-eclampsia (9.2% vs. 5.3%, p<0.0001) and gestational diabetes mellitus (10.3% vs. 7.9%, p<0.0001) when compared to controls. This was also seen among pregnancies with SVT with pre-eclampsia (8.0% vs. 5.3%, p<0.0001) and gestational diabetes mellitus (9.9% vs. 7.9%, p<0.0001).

**Table 3.**
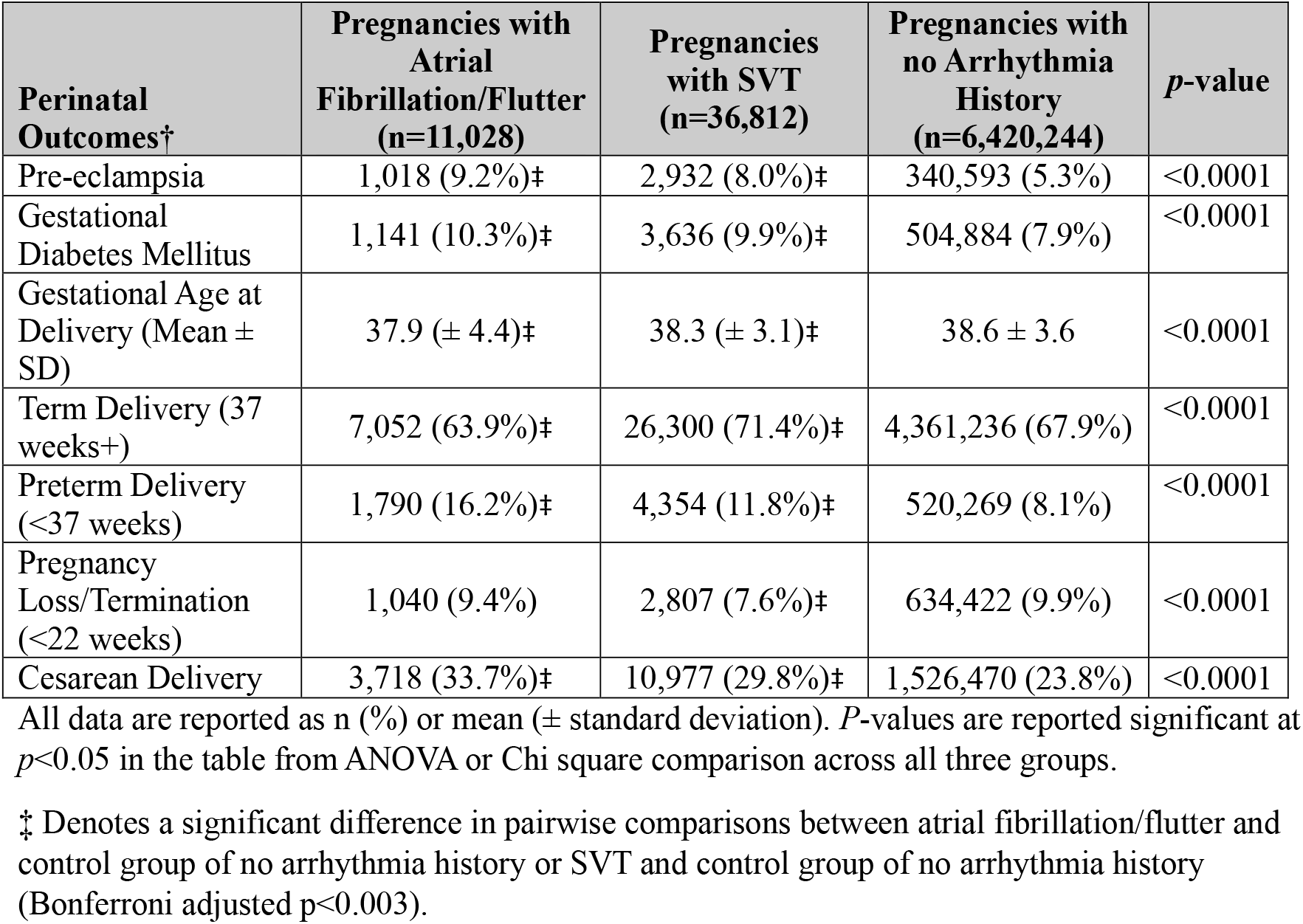
Perinatal Outcomes of Pregnant Patients with Cardiac Arrhythmias.

| **Perinatal Outcomes†** | **Pregnancies with Atrial Fibrillation/Flutter (n=11,028)** | **Pregnancies with SVT (n=36,812)** | **Pregnancies with no Arrhythmia History (n=6,420,244)** | ***p*-value** |
| --- | --- | --- | --- | --- |
| Pre-eclampsia | 1,018 (9.2%)‡ | 2,932 (8.0%)‡ | 340,593 (5.3%) | <0.0001 |
| Gestational Diabetes Mellitus | 1,141 (10.3%)‡ | 3,636 (9.9%)‡ | 504,884 (7.9%) | <0.0001 |
| Gestational Age at Delivery (Mean ± SD) | 37.9 (± 4.4)‡ | 38.3 (± 3.1)‡ | 38.6 ± 3.6 | <0.0001 |
| Term Delivery (37 weeks+) | 7,052 (63.9%)‡ | 26,300 (71.4%)‡ | 4,361,236 (67.9%) | <0.0001 |
| Preterm Delivery (<37 weeks) | 1,790 (16.2%)‡ | 4,354 (11.8%)‡ | 520,269 (8.1%) | <0.0001 |
| Pregnancy Loss/Termination (<22 weeks) | 1,040 (9.4%) | 2,807 (7.6%)‡ | 634,422 (9.9%) | <0.0001 |
| Cesarean Delivery | 3,718 (33.7%)‡ | 10,977 (29.8%)‡ | 1,526,470 (23.8%) | <0.0001 |

All data are reported as n (%) or mean ( ± standard deviation). *P*-values are reported significant at $p<0.05$ in the table from ANOVA or Chi square comparison across all three groups.
‡ Denotes a significant difference in pairwise comparisons between atrial fibrillation/flutter and control group of no arrhythmia history or SVT and control group of no arrhythmia history (Bonferroni adjusted $p<0.003$ ).

## Discussion

Through a large, contemporary database, this study showed that both AF/AFL and SVT are associated with adverse cardiac and perinatal outcomes, but that AF/AFL is associated with the highest risk of these outcomes including preterm birth, maternal stroke, and heart failure in pregnancy.

Most significantly, our research highlights that AF/AFL carries a high risk of adverse outcomes including stroke in pregnancy when compared to SVT or pregnancies not impacted by arrhythmias. The stroke risk is higher than what is seen in other literature, however our sample size is much larger than other published studies and may be more representative of the pregnant population than single centers.^9,10^ While the CHA_2_DS_2_-VASc score can be used to estimate stroke risk, it may not be as reliable in a pregnant population.^11^ Pregnancy is a known pro-thrombotic state, and AF/AFL significantly increases this risk. Given many pregnant patients may be young and otherwise healthy, they may have a lower CHA_2_DS_2_-VASc score and thereby not meet criteria for anticoagulation leading to decreased utilization. However, there were high rates of beta blocker and calcium channel blocker use in this population, which may reflect increased rates of chronic hypertension or hypertensive disorders of pregnancy. While hypertension and female sex would be reflected in the CHA_2_DS_2_-VASc score, a pregnancy specific risk score considering congenital heart disease, gestational diabetes, other hypertensive disorders of pregnancy, and obesity should be developed to improve clinical decision making in pregnancy and postpartum.

Conversely, the risks of anticoagulation in pregnancy must also be considered. While enoxaparin is well studied and can be administered in pregnancy, timing of anticoagulation can impact the ability to obtain neuraxial anesthesia and impact delivery.^12^ Other agents like warfarin or other direct oral anticoagulants are teratogenic or have unknown fetal impacts. The risks and benefits of anticoagulation in pregnancy in the setting of AF/AFL should be discussed with patients given the increased risk of stroke seen in this population. It may be reasonable to offer all pregnant patients with AF/AFL treatment with anticoagulation regardless of their CHA_2_DS_2_-VASc score if an additional pregnancy-specific risk factor for thromboembolism is present, given that a 4% stroke risk in one year corresponds to a high-risk CHA2DS2-VASc score of 4 or greater.^13^ While over 50% of the patients with AF/AFL were on a form of an anticoagulant medication during the pregnancy, limitations in Epic Cosmos prevent us from determining the dosing (prophylactic or therapeutic), specific indication, or duration/timing of use.

The World Health Organization classification system of cardiac disease in pregnancy categorizes most cardiac arrhythmias as WHO class II, which is deemed as low to moderate risk.^14^ Based on the findings of this study, AF/AFL should be considered a higher risk cardiac condition given that over 10% of pregnant patients experienced heart failure with AF and nearly 4% suffered a stroke. Pregnant patients with SVT had a lower rate of cardiac events, which is reassuring for pregnancy management and supports the appropriateness of a WHO class II risk categorization.

The adverse perinatal outcome of an increased risk of preterm birth with AF is consistent with other literature on cardiac arrhythmias.^15^ The increase in preterm deliveries is likely to be iatrogenic in the setting of maternal hemodynamic instability requiring delivery, difficulty with rate or rhythm control in advancing gestation, or the increased presence of comorbid conditions like hypertensive disorders of pregnancy necessitating preterm delivery.

Future research should aim to further delineate the risks of different types of cardiac arrhythmias in pregnancy and the postpartum period to better allow for patient counseling of risks. Studies detailing the timing of adverse cardiac events in pregnancy or the postpartum period will be crucial in guiding clinicians in the creation of antepartum, intrapartum, and postpartum care plans.

### Strengths and Limitations

The strengths of this study include the large sample size of the populations assessed and differentiation between two common arrhythmias seen in pregnancy, given preexisting data is limited in pregnant populations. Prior data often groups pregnant patients with any cardiac arrhythmias together due to overall small sample sizes, which may mask the risks of certain arrythmias. The limitations of this study include a reliance on ICD-10 codes for diagnoses in pregnancy and the inability to confirm the temporal relation of the diagnosis codes during the pregnancy (i.e. pregnancy trimester vs. postpartum period). Additionally, we do not have access to individual level data in Epic Cosmos, which limits our ability to perform multivariate adjusted logistic regressions. While our study is limited to unadjusted analyses due to the presence of only aggregate level data, it highlights important trends related to the risks of AF/AFL and stroke that warrants further investigation.

### Conclusion

In conclusion, cardiac arrhythmias are associated with adverse outcomes in pregnancy, with AF predisposing pregnant patients to a higher risk of adverse cardiac and pregnancy outcomes than other cardiac arrhythmias like SVT. Future studies should aim to examine pregnancy-specific AF/AFL management plans and the timing of adverse events in pregnancy to help further inform clinical guideline development and clinicians caring for this high-risk population.

## Data Availability

The data is available through Epic Cosmos and can be shared at request. All data is deidentified.

